# A transdiagnostic systematic review and meta-analysis of ketamine’s anxiolytic effects

**DOI:** 10.1101/2022.12.09.22283264

**Authors:** Hannah Hartland, Kimia Mahdavi, Luke A Jelen, Rebecca Strawbridge, Allan H Young, Laith Alexander

## Abstract

**Background:** Subanaesthetic doses of ketamine may be effective in treating symptoms of anxiety, but the time profile of ketamine’s anxiolytic effect is ill-defined. This systematic review and meta-analysis of randomised controlled trials investigated the anxiolytic effect of ketamine at different time points across a range of clinical contexts.

**Methods:** Electronic databases were searched to capture randomised control trials measuring the anxiolytic effects of ketamine across a range of clinical settings including mood disorders, anxiety disorders and chronic pain. Meta-analyses were conducted using a random-effects model. The correlations between (1) improvements in mean anxiety and depression scores, and (2) peak dissociation and improvements in mean anxiety scores were also assessed.

**Results:** 14 studies met inclusion criteria. Risk of bias was high in 11 studies. Ketamine significantly reduced anxiety scores compared to placebo at the acute (<12 hours; SMD: - 1.07 [95% CI: -1.68, -0.46], p < 0.001), subacute (24 hours; SMD: -0.43 [95% CI: -0.65, -0.22], p < 0.001) and sustained (7-14 days; SMD: -0.43 [95% CI: -0.65, -0.20], p < 0.001) time points. Exploratory analyses revealed improvements in anxiety and depression symptoms correlated at both subacute (R^2^ = 0.621, *p* = 0.035) and sustained time points (R^2^ = 0.773, *p* = 0.021). The relationship between peak dissociation and improvement in anxiety was not significant.

**Conclusions:** Ketamine appears to offer rapid and sustained relief from anxiety symptoms across a range of clinical settings, with anxiolytic effects occurring within the first 12 hours of administration and remaining effective for one to two weeks. Future studies with improved blinding could explore ketamine maintenance therapy for anxiety.

## Introduction

Anxiety disorders are common, with a lifetime prevalence of 33.7% in United States adults (Bandelow & Michaelis, 2015). These disorders share a common factor of fear and anxiety which impair daily functioning but differ in the situations or objects catalysing the schema (American Psychiatric Association, 2013). Significant anxiety symptoms are seen in a range of clinical settings, including major depressive disorder (MDD) (Drysdale et al., 2017; Park & Kim, 2020), chronic pain (McWilliams et al., 2003), advanced cancer (Roth & Massie, 2007), and palliative care (Kelly et al., 2006; Kozlov et al., 2019; Neel et al., 2015).

Current treatments for pathological anxiety include selective serotonin reuptake inhibitors (SSRIs), but these have a slow onset of action and can paradoxically worsen anxiety upon initiation (Cassano et al., 2002; Nutt, 2005). Benzodiazepines provide rapid relief of acute anxiety symptoms, but are associated with tolerance and dependence (Bystritsky, 2006; Cassano et al., 2002). Given these issues and the prevalence of treatment-resistant anxiety (Bystritsky, 2006), there is a pressing need for the development of novel rapidly acting anxiolytic agents with the potential for intermediate to long-term administration.

One avenue which may meet this need comes in the form of the N-methyl-D-aspartic acid (NMDA) receptor antagonist, ketamine. Ketamine is an effective glutamate-based antidepressant (Marcantoni et al., 2020), but evidence has also emerged supporting its anxiolytic effects. In patients with social anxiety disorder (SAD) (Glue et al., 2018; Taylor et al., 2018), obsessive-compulsive disorder (OCD) (Bandeira et al., 2022), post-traumatic stress disorder (PTSD) (Liriano et al., 2019), and treatment-refractory anxiety (Tully et al., 2022), ketamine may have fast-acting anxiolytic effects lasting approximately one to two weeks after a single dose.

Several review articles have inferred ketamine’s utility in the treatment of anxiety disorders and have supported this suggestion (Banov et al., 2020; Tully et al., 2022; Whittaker et al., 2021). However, these reviews did not examine the time course of ketamine’s action, and included open-label studies, uncontrolled trials, case series, and other lower quality studies, limiting the conclusions that can be drawn about ketamine’s anxiolytic effects compared to placebo. Furthermore, the transdiagnostic nature of anxiety symptoms, as emphasised by the negative valence system in the *Research Domain Criteria* (Insel et al., 2010) and the fear and distress subfactors in the *Hierarchical Taxonomy of Psychopathology* (Kotov et al., 2017), means that ketamine may prove to be useful in a range of clinical settings associated with distressing anxiety symptoms and this has yet to be explored.

The purpose of this systematic review and meta-analysis is to examine the effect of ketamine on symptoms of anxiety at several time points, through synthesizing the findings of blinded, randomised, placebo-controlled trials (RCTs). Our principal analyses explored ketamine’s anxiolytic effects acutely (< 12 hours), subacutely (at 24 hours) and at a sustained time point (7-14 days), incorporating data from RCTs measuring anxiety symptoms in anxiety disorders, depression & other mood disorders, chronic pain, and palliative care settings.

## Methods

This review was carried out in accordance with PRISMA guidance. We registered the protocol for this review with the PROSPERO International prospective register of systematic reviews (CRD42022303070; [URL](https://www.crd.york.ac.uk/prospero/display_record.php?RecordID=303070)). In addition to the protocol described on PROSPERO, we collected depression and dissociation data and correlated these against changes in anxiety scores as described in **Data extraction and analysis**.

### Inclusion and exclusion criteria

Our inclusion criteria were:

- **Study type**: single or double blinded RCT written in English (including crossover trials)
- **Population**: adult human patients suffering from anxiety disorders of any type (including post-traumatic stress disorder [PTSD] and obsessive-compulsive disorder [OCD]) or in whom anxiety symptoms were measured in the context of mood disorders, chronic pain or palliative care
- **Intervention**: subanaesthetic doses of either racemic ketamine, *S*-ketamine or *R*-ketamine, administered via intravenous, intranasal, oral, subcutaneous, intramuscular or sublingual routes
- **Control**: an active or inactive placebo comparator
- **Outcome**: a primary or secondary outcome relating to anxiety

We excluded:

- Animal studies
- Non-RCT studies or where no party was blinded to treatment allocation
- Any studies where ketamine was used at anaesthetic doses or in the context of surgery/anaesthesia
- Any studies where ketamine was administered in the context of a medical or surgical emergency
- Studies with child or adolescent subjects

### Search strategy

EMBASE (Ovid), MEDLINE (Ovid) and APA PsycINFO (Ovid) were systematically searched (February 2022) using keywords and medical subject headings relating to ketamine and anxiety: **ketamine** AND [(**Anxiety** OR **Test Anxiety** OR **Anxiety Disorders** OR **Test Anxiety Scale**) OR **Obsessive-Compulsive Disorder** OR **Social Phobia** OR **Phobic Disorders** OR **Agoraphobia** OR **Post-Traumatic Stress Disorders** OR **Separation Anxiety** OR (**Panic Disorder** OR **Panic**) OR **Mutism** OR (**GAD-7** OR **Patient Health Questionnaire**) OR (**Palliative Care** OR **“Hospice and Palliative Care Nursing”** OR **Palliative Medicine**)] AND (**Randomized Controlled Trial** OR **Clinical Trial**) NOT **Child**. Additionally, the first 200 findings based on relevance of a Google Scholar search (February 2022) using keywords (**ketamine** AND **anxiety**) were examined.

### Article screening and assessment for eligibility

Any duplicate titles and abstracts generated from the search were removed using Syras systematic review screening software (Scipilot Pty. Ltd., Sydney, Australia). The remaining articles were screened for potential relevance and eligibility according to pre-specified inclusion and exclusion criteria by three independent researchers, with others’ ratings masked from their own.

### Risk of Bias

To determine reliability and transparency of the studies, a risk of bias assessment was completed for each included RCT using the Cochrane risk-of-bias assessment tool for randomized trials (Higgins et al., 2011). The Cochrane tool explores bias over five domains: selection bias, performance bias, detection bias, attrition bias, and reporting bias. There is an additional domain for crossover trials which explores any carryover bias. Each study was judged across all domains as well as given an overall rating. The risk of bias assessment was completed independently by three reviewers, ensuring each study was double rated by two different reviewers. Disagreements were discussed and resolved through consensus.

### Data extraction and analysis

Descriptive data were extracted by three independent reviewers and organised into a table (**Table 1**) including the type of study (single or double-blinded RCT), population (disorder, number and whether medication-free) studied, intervention used (type and dose(s) of ketamine, route of administration, and frequency of dosing if applicable), control used (details of the inactive or active placebo), and outcome measured (anxiety scale used; anxiety scores at time points measured).

**Table 1.**
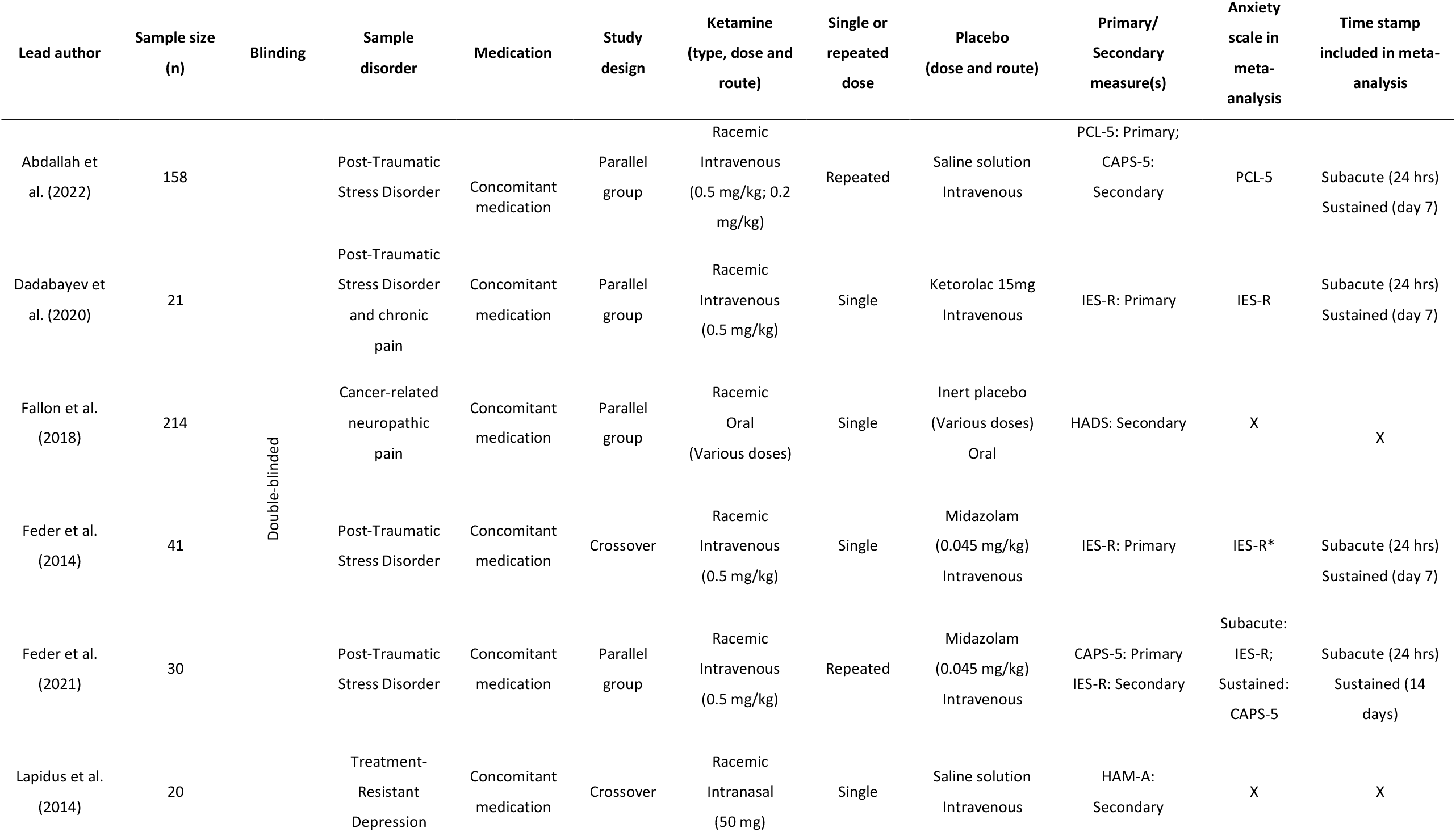
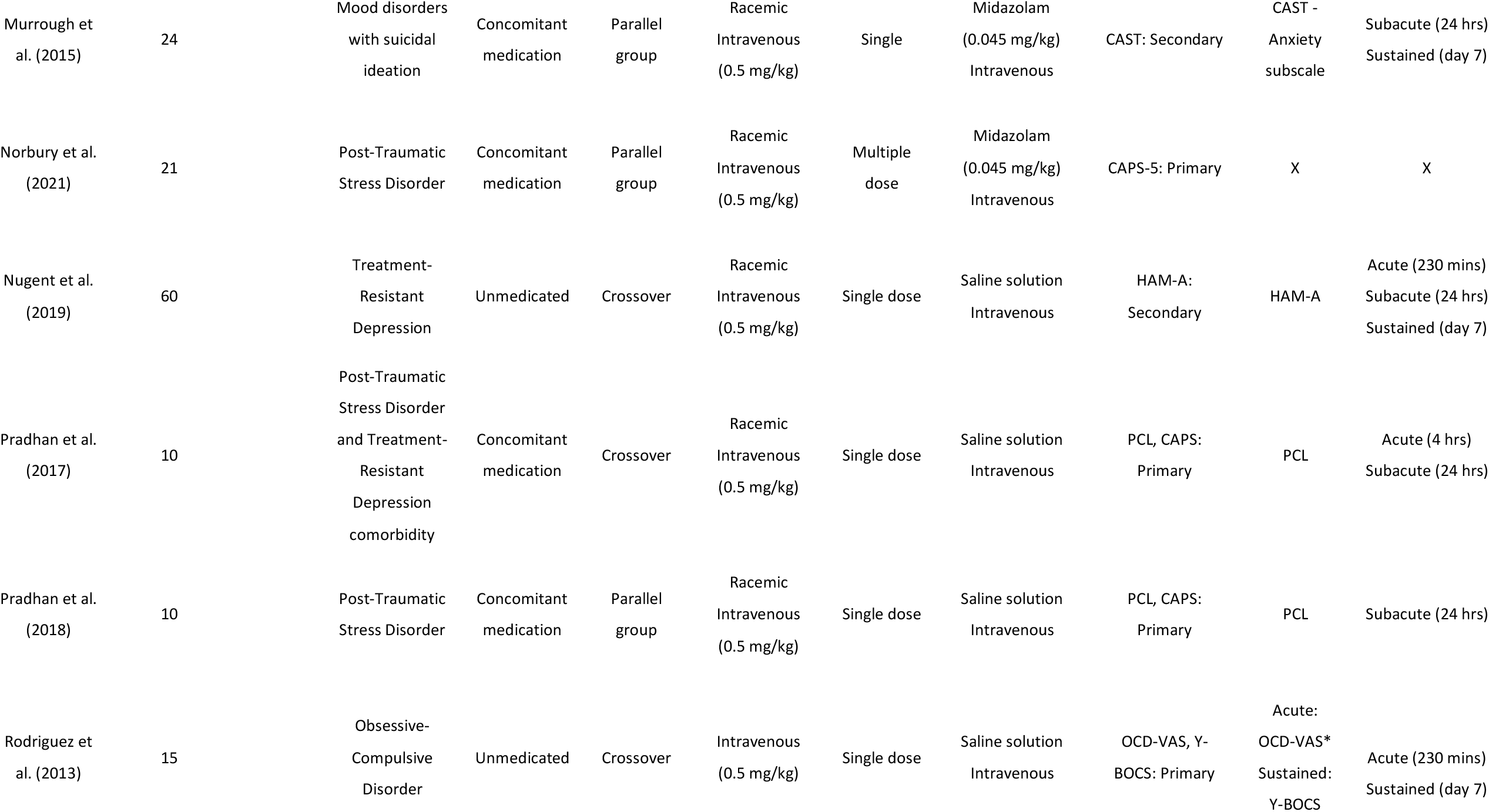
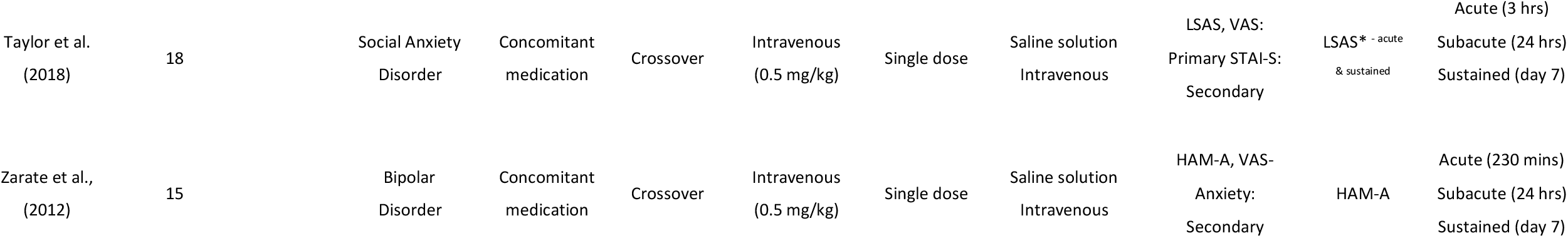
Study characteristics. Abbreviations: PCL, Posttraumatic Stress Disorder Checklist; PCL-5, Posttraumatic Stress Disorder Checklist for DSM-5; CAPS, Clinical Administered PTSD Scale; CAPS-5, Clinically Administered Posttraumatic Stress Disorder Scale for DSM-5 ;IES-R, Impact of Events Scale-Revised; HAM-A, Hamilton Anxiety Rating Scale; CAST, Concise Associated Symptoms Tracking scale ; Y-BOCS, Yale-Brown Obsessive Compulsive Scale; OCD-VAS, Obsessive Compulsive Disorder-Visual Analogue Scale ; Anxiety-VAS; Anxiety-Visual Analogue Scale; LSAS, Liebowitz Social Anxiety Scale; STAI-S, State-Trait Anxiety Subscale. * indicates that the value included in meta-analysis was estimated.

| Lead author | Sample size (n) | Blinding | Sample disorder | Medication | Study design | Ketamine (type, dose and route) | Single or repeated dose | Placebo (dose and route) | Primary/ Secondary measure(s) | Anxiety scale in meta-analysis | Time stamp included in meta-analysis |
| --- | --- | --- | --- | --- | --- | --- | --- | --- | --- | --- | --- |
| Abdallah et al. (2022) | 158 | Double-blinded | Post-Traumatic Stress Disorder | Concomitant medication | Parallel group | Racemic Intravenous (0.5 mg/kg; 0.2 mg/kg) | Repeated | Saline solution Intravenous | PCL-5: Primary; CAPS-5: Secondary | PCL-5 | Subacute (24 hrs) Sustained (day 7) |
| Dadabayev et al. (2020) | 21 |  | Post-Traumatic Stress Disorder and chronic pain | Concomitant medication | Parallel group | Racemic Intravenous (0.5 mg/kg) | Single | Ketorolac 15mg Intravenous | IES-R: Primary | IES-R | Subacute (24 hrs) Sustained (day 7) |
| Fallon et al. (2018) | 214 |  | Cancer-related neuropathic pain | Concomitant medication | Parallel group | Racemic Oral (Various doses) | Single | Inert placebo (Various doses) Oral | HADS: Secondary | X | X |
| Feder et al. (2014) | 41 |  | Post-Traumatic Stress Disorder | Concomitant medication | Crossover | Racemic Intravenous (0.5 mg/kg) | Single | Midazolam (0.045 mg/kg) Intravenous | IES-R: Primary | IES-R* | Subacute (24 hrs) Sustained (day 7) |
| Feder et al. (2021) | 30 |  | Post-Traumatic Stress Disorder | Concomitant medication | Parallel group | Racemic Intravenous (0.5 mg/kg) | Repeated | Midazolam (0.045 mg/kg) Intravenous | CAPS-5: Primary IES-R: Secondary | Subacute: IES-R; Sustained: CAPS-5 | Subacute (24 hrs) Sustained (14 days) |
| Lapidus et al. (2014) | 20 |  | Treatment-Resistant Depression | Concomitant medication | Crossover | Racemic Intranasal (50 mg) | Single | Saline solution Intravenous | HAM-A: Secondary | X | X |

**Table 1.** Study characteristics.
|  |  |  |  |  |  |  |  |  |  |  |
| --- | --- | --- | --- | --- | --- | --- | --- | --- | --- | --- |
| Murrough et al. (2015) | 24 | Mood disorders with suicidal ideation | Concomitant medication | Parallel group | Racemic Intravenous (0.5 mg/kg) | Single | Midazolam (0.045 mg/kg) Intravenous | CAST: Secondary | CAST - Anxiety subscale | Subacute (24 hrs)<br>Sustained (day 7) |
| Norbury et al. (2021) | 21 | Post-Traumatic Stress Disorder | Concomitant medication | Parallel group | Racemic Intravenous (0.5 mg/kg) | Multiple dose | Midazolam (0.045 mg/kg) Intravenous | CAPS-5: Primary | X | X |
| Nugent et al. (2019) | 60 | Treatment-Resistant Depression | Unmedicated | Crossover | Racemic Intravenous (0.5 mg/kg) | Single dose | Saline solution Intravenous | HAM-A: Secondary | HAM-A | Acute (230 mins)<br>Subacute (24 hrs)<br>Sustained (day 7) |
| Pradhan et al. (2017) | 10 | Post-Traumatic Stress Disorder and Treatment-Resistant Depression comorbidity | Concomitant medication | Crossover | Racemic Intravenous (0.5 mg/kg) | Single dose | Saline solution Intravenous | PCL, CAPS: Primary | PCL | Acute (4 hrs)<br>Subacute (24 hrs) |
| Pradhan et al. (2018) | 10 | Post-Traumatic Stress Disorder | Concomitant medication | Parallel group | Racemic Intravenous (0.5 mg/kg) | Single dose | Saline solution Intravenous | PCL, CAPS: Primary | PCL | Subacute (24 hrs) |
| Rodriguez et al. (2013) | 15 | Obsessive-Compulsive Disorder | Unmedicated | Crossover | Intravenous (0.5 mg/kg) | Single dose | Saline solution Intravenous | OCD-VAS, Y-BOCS: Primary | Acute: OCD-VAS*<br>Sustained: Y-BOCS | Acute (230 mins)<br>Sustained (day 7) |
| Taylor et al.<br>(2018) | 18 | Social Anxiety<br>Disorder | Concomitant<br>medication | Crossover | Intravenous<br>(0.5 mg/kg) | Single dose | Saline solution<br>Intravenous | LSAS, VAS:<br>Primary STAI-S:<br>Secondary | LSAS* - acute<br>& sustained | Acute (3 hrs)<br>Subacute (24 hrs)<br>Sustained (day 7) |
| Zarate et al.,<br>(2012) | 15 | Bipolar<br>Disorder | Concomitant<br>medication | Crossover | Intravenous<br>(0.5 mg/kg) | Single dose | Saline solution<br>Intravenous | HAM-A, VAS-Anxiety:<br>Secondary | HAM-A | Acute (230 mins)<br>Subacute (24 hrs)<br>Sustained (day 7) |

Numerical data were extracted either directly from published papers (when available), from direct communication with authors, or from estimating graph values if needed. Extracted numerical data were then compiled using Review Manager (RevMan version 5.4), and RevMan was used to generate forest plots of the standard mean differences (SMDs) in anxiety scores between groups receiving ketamine *vs*. placebo. Separate analyses were carried out on three time points: less than 12 hours post-administration (acute); 24 hours post-administration (subacute); and 7-14 days post-administration (sustained). A time point beyond 14 days was also qualitatively assessed. To maximise comparability, when a study had multiple time points which fell under one of our pre-specified ranges, the modal time point that was available across studies was included (unless a study only had data available at a different time point). In the case of multiple anxiety outcomes, the primary outcome as defined by the study was used. In crossover trials where a carryover effect was identified by the authors, data from the first arm only were included in the meta-analysis.

Data were pooled across studies to conduct exploratory analyses of the correlation between improvements in anxiety scores post-ketamine and (1) improvements in depression scores post-ketamine and (2) peak Clinician-Administered Dissociative States Scale (CADSS) scores using linear regressions in R (version 3.5.3).

## Results

Results for the systematic review and meta-analysis article search are summarised in the PRISMA flowchart (**Figure 1**). The combined searches generated 4647 records, leaving 4515 once duplicates were removed. After initial screening, 309 articles were included in the full-text review. Of these, 295 were deemed ineligible, and 14 RCTs were included in the qualitative systematic review. Due to missing and inaccessible data, 3 were excluded from quantitative analysis, meaning 11 articles were included in the meta-analysis.

**Figure 1.**
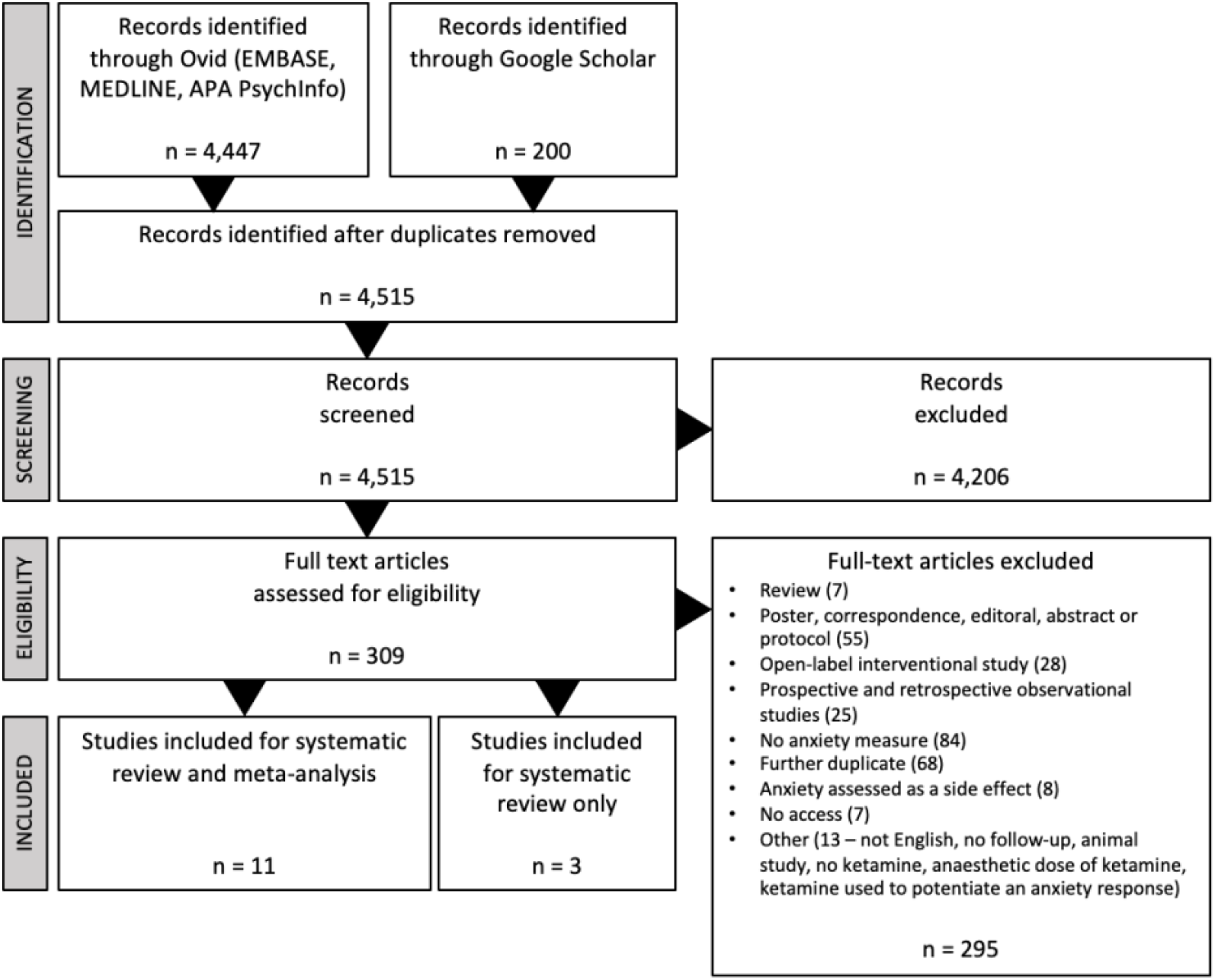
PRISMA flow-chart.

### Study characteristics and risk of bias

Information about each study, including design, sample sizes, ketamine dosing, control dosing, and outcome measures is reported in **Table 1**. Results of the Cochrane risk of bias analysis revealed that all but two studies had an overall rating of some concerns or high risk of bias (**Figure 2**). The most common domains of concern were deviations from intended interventions, selective reporting of results and carryover effects in two of the seven studies with crossover designs.

**Figure 2.**
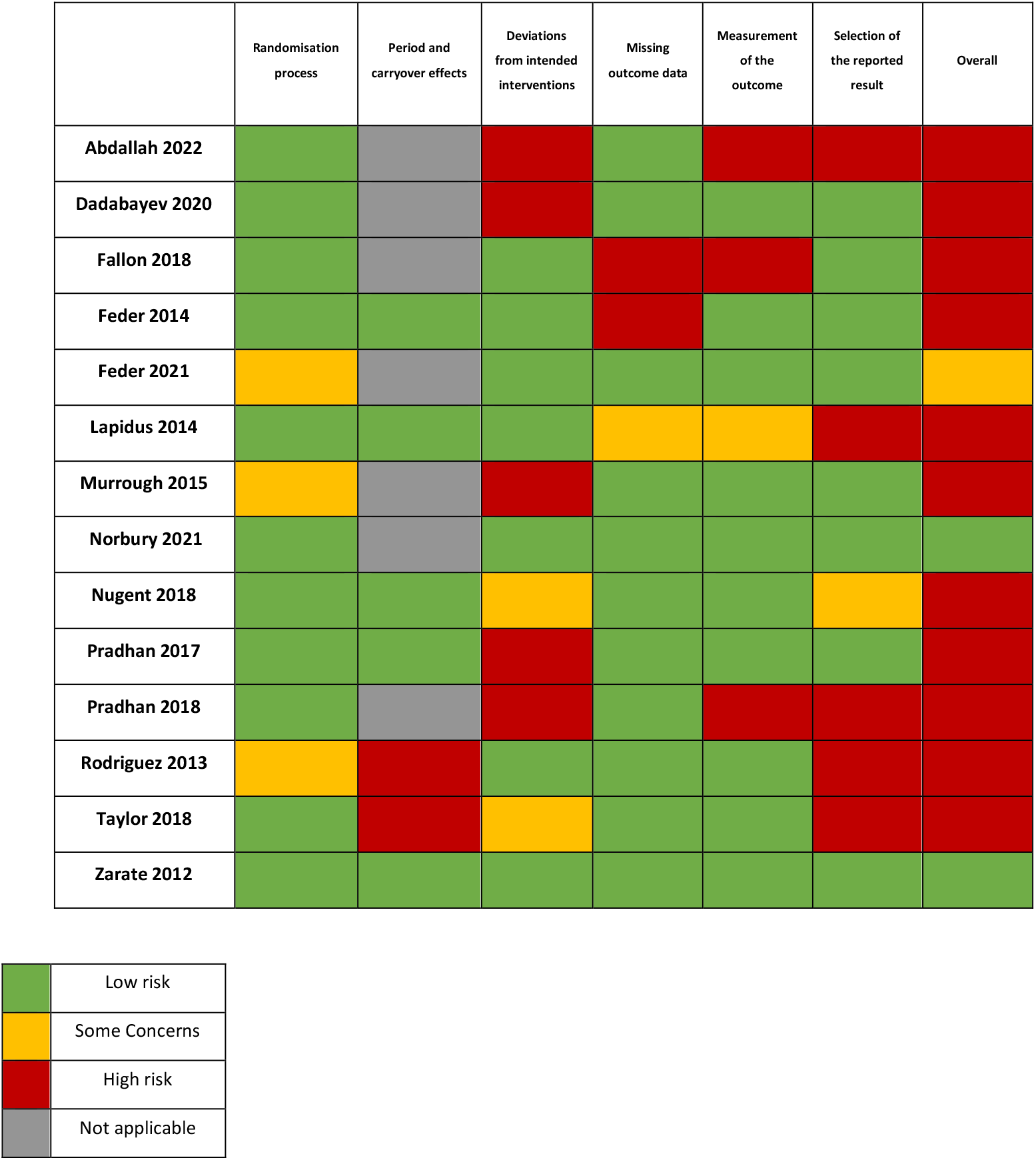
Cochrane risk of bias table for included studies (*k* = 14).

The subsequent sections present analyses divided into ketamine’s acute response (less than 12 hours; *k* = 6), subacute response (24 hours; *k* = 10), sustained response (7-14 days; *k* = 9), and responses beyond 14 days (*k* = 4). For a detailed summary of significant and non-significant findings of each study, see **Supplementary table 1**.

### Acute (<12 hours)

Seven studies measured anxiety at one or more time points <12 hours after ketamine administration. Two of these studies did not report their findings (Lapidus et al., 2014; Pradhan et al., 2018). Of the rest, three reported that ketamine reduced anxiety significantly compared to placebo at at least one time point <12 hours (Nugent et al., 2018; Rodriguez et al., 2013; Zarate et al., 2012), and two reported non-significance compared to placebo (Pradhan et al., 2017; Taylor et al., 2018).

All three significant studies used intravenous ketamine. Zarate et al. (2012) (*n* = 15) and Nugent et al. (2018) (*n* = 35) explored anxiolytic effects in mood disorder participants, and Rodriguez et al. (2013) (*n* = 15) explored effects in patients with OCD. Zarate et al. (2012) found significant improvements in the Visual Analogue Scale for anxiety (VAS-Anxiety; Aitken, 1969) starting at 40 minutes post-infusion, which remained significant at 80 minutes, 110 minutes and 230 minutes. Nugent et al. (2018) found significant improvements in Hamilton Anxiety Scale (HAM-A; Hamilton, 1959) scores at 230 minutes, but not 40 minutes. Similarly, Rodriguez et al. (2013) found significant improvements in the Visual Analogue Scale for OCD (OCD-VAS; Rodriguez et al., 2011) scores at 230 minutes, but not 90 minutes and 110 minutes.

For the meta-analysis, group-level data were obtained for all five studies with reported data taken between three- and four-hours post-administration (**Figure 3A**). This meta-analysis included 69 patients who received ketamine and 63 who received placebo. Standardised mean differences (SMDs) were calculated for each study, as well as an overall SMD for the meta-analysis, which was significant in favour of ketamine compared to placebo (SMD: - 1.07, 95% CI: [-1.68, -0.46], *p* < 0.01). There was moderate heterogeneity among the studies, but this was not statistically significant (I^2^ = 51%, *p* = 0.08).

**Figure 3.**
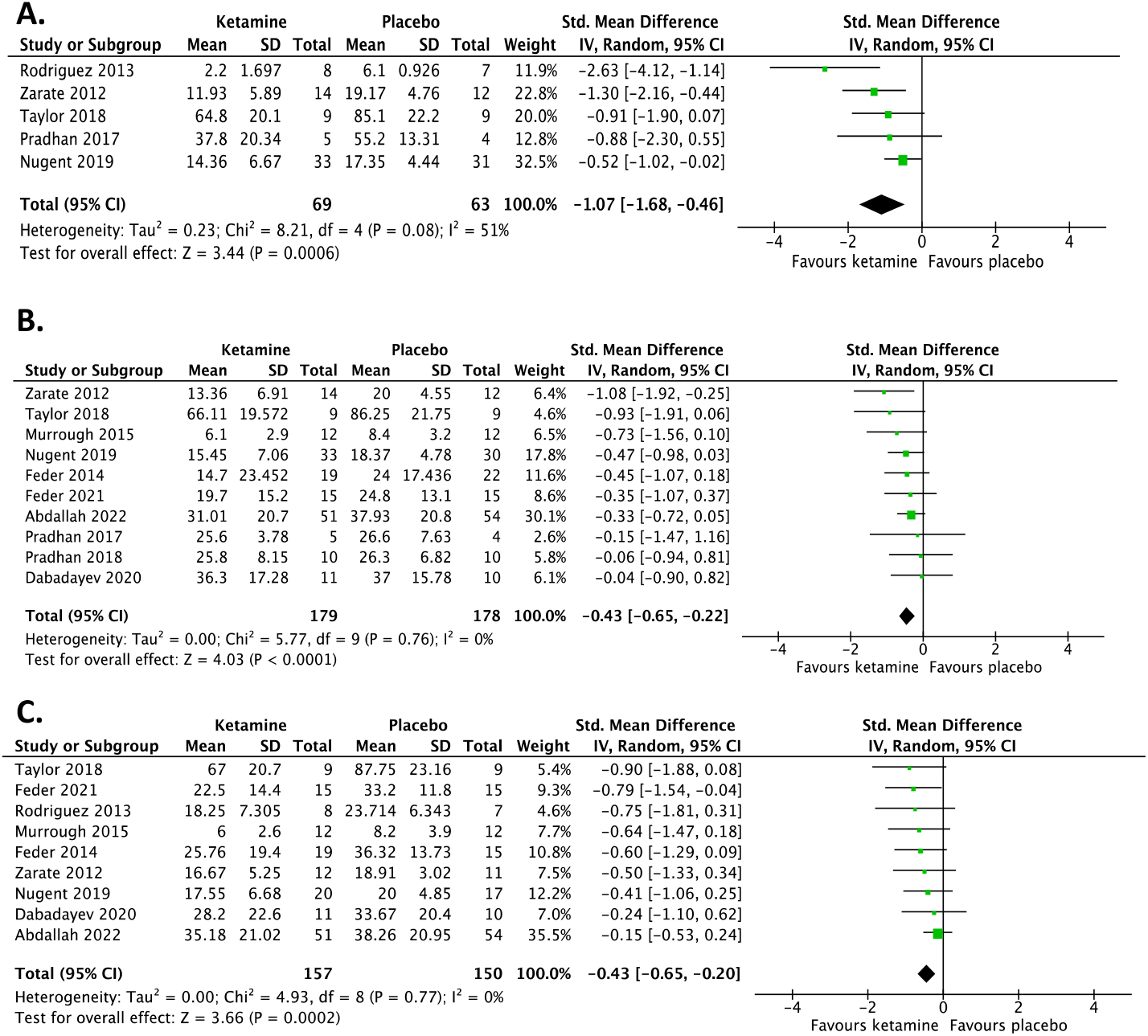
Forest plots. Left favours ketamine (indicates ketamine reduced anxiety scores compared to control). Error bars represent 95% confidence intervals. (**A**) Acute time point (<12 hours). (**B**) Subacute time point (24 hours). (**C**) Sustained time point (7-14 days).

### Subacute (24 hours)

Most studies included in this review reported findings at 24 hours post-ketamine administration (*k* = 10). Of these studies, four reported a significant effect of ketamine in reducing anxiety symptoms compared to placebo: three using the HAM-A scale in mood disorder patients and one using the Impact of Event Scale–Revised (IES-R; Weiss & Marmar, 1997) in PTSD patients (Feder et al., 2014; Lapidus et al., 2014; Nugent et al., 2019; Zarate et al., 2012). All four studies used a crossover design.

Respectively, Lapidus et al. (2014) (*n* = 20) and Nugent et al. (2019) (*n* = 60) found intranasal and intravenous ketamine treatments to be more effective in reducing HAM-A scores in MDD patients than placebo at 24 hours post-administration. Similarly, Zarate et al. (2012) (*n* = 15) found intravenous ketamine to be superior to placebo at symptom reduction using this scale at 24 hours in bipolar disorder patients. Lastly, Feder et al. (2014) (*n* = 41) reported significant drug group differences at 24 hours on the IES-R when comparing the efficacy of 0.5 mg/kg ketamine with an active placebo, midazolam (0.045 mg/kg) in 41 patients with chronic PTSD.

Results of the meta-analysis of 10 studies with available group-level data (**Figure 3B**) showed that there was a significant difference in anxiety scores between the ketamine (*n* = 179) and placebo (*n* = 178) groups (SMD: -0.43, 95% CI: [-0.65, - 0.22], *p* < 0.001). No significant heterogeneity was reported (I^2^ = 0%, *p* = 0.76).

### Sustained (7-14 days)

There were mixed findings from the nine studies which looked at time points between seven- and 14-days post-infusion – four reported significant findings (Feder et al., 2021; Rodriguez et al., 2013; Zarate et al., 2012; Taylor et al., 2018), and three reported no significance (Abdallah et al., 2022; Feder et al., 2014; Nugent et al., 2018).

Feder et al. (2021) (*n* = 30) used scores from the Clinician-Administered PTSD Scale for DSM-5 (CAPS-5; Weathers et al., 2018) to assess symptom severity one week into treatment with multiple doses of intravenous ketamine (measured after fourth infusion) and at the end of treatment. Analysis revealed significantly lower total scores in the ketamine group compared to the midazolam group at one week post-first infusion, which was sustained at two-weeks post-first infusion. Two studies with significant findings recruited participants with an anxiety disorder diagnosis. Rodriguez et al. (2013) (*n* = 15) found that those who received ketamine reported significantly lower means on the OCD-VAS at seven days post-infusion. Meanwhile, Taylor and colleagues (2018) (*n* = 18) found that, starting at 10 days post-infusion, participants with SAD receiving ketamine demonstrated significantly greater reductions in overall Liebowitz Social Anxiety Scale (LSAS; Heimberg et al., 1999) scores compared to those receiving placebo.

Both Murrough et al. (2015) and Zarate et al. (2012) explored ketamine’s efficacy in patients diagnosed with mood disorders. In patients with mood disorders and clinically significant suicidal ideation, Murrough and colleagues (2015) (*n* = 24) found that mean Concise Associated Symptoms Tracking scale (CAST; Trivedi et al., 2011) subscale scores pertaining to anxiety (irritability, anxiety and panic) were numerically lower in the ketamine compared to midazolam group on each subscale at seven days, but the researchers did not analyse the CAST subscale scores statistically. Meanwhile, Zarate et al. (2012) reported significantly lower scores in subjects who had received ketamine as opposed to placebo on the VAS-Anxiety scale at days seven and 14 post-ketamine administration.

The meta-analysis included nine studies for which data were available (**Figure 3C**) and included 157 patients who had received ketamine and 150 patients who had received placebo. Analysis revealed that mean anxiety scores were significantly lower in the ketamine group compared to those in the placebo group (SMD: -0.43 [95% CI: -0.65, -0.20], *p* < 0.01). There was no significant heterogeneity among studies (I^2^ = 0%, *p* < 0.001).

### Effects beyond 14 days

Of the four studies which explored the efficacy of ketamine beyond two weeks, two studies reported significant treatment group differences (Norbury et al., 2021; Pradhan et al., 2018), whilst two reported non-significance (Pradhan et al., 2017; Fallon et al., 2018).

A repeated dose study carried out by Norbury and colleagues (2021) found CAPS-5 scores to be significantly lower at 16 days post-first dose in PTSD patients who had received ketamine than those who had received midazolam. Results revealed a significant session-by-drug interaction on CAPS-5 scores (*F*_1,57_ = 6.58, *p* = 0.013). Also exploring a prolonged treatment response were the Pradhan studies (Pradhan et al., 2017 & 2018), who measured Clinical Administered PTSD Scale (CAPS; Blake et al., 1995) and PTSD Checklist for DSM-IV (PCL; Weathers et al., 1993) scores weekly until relapse. In the 2017 study, patients receiving ketamine had a more sustained response (33 ± 22.98 days) than those who received placebo (25 ± 16.8 days), though this difference was non-significant (*p* = 0.545). In their 2018, the difference in length of response demonstrated a similar pattern (34.44 ± 19.12 days in the ketamine group and 16.50 ± 11.39 in the placebo group), however the difference was significant (*p* = 0.022).

### Correlation between improvement in anxiety and depression scores

Seven studies provided depression and anxiety data at the subacute time point and six studies provided this data at sustained time point, leading to 182 and 175 participants included in each analysis respectively. A linear regression analysis was conducted and found that there was a significant correlation between percentage improvement in anxiety and depression scores at the subacute time point (R^2^ = 0.621, *p* = 0.035, **Figure 4A**) and at the sustained time point (R^2^ = 0.773, *p* = 0.021; **Figure 4B**).

**Figure 4.**
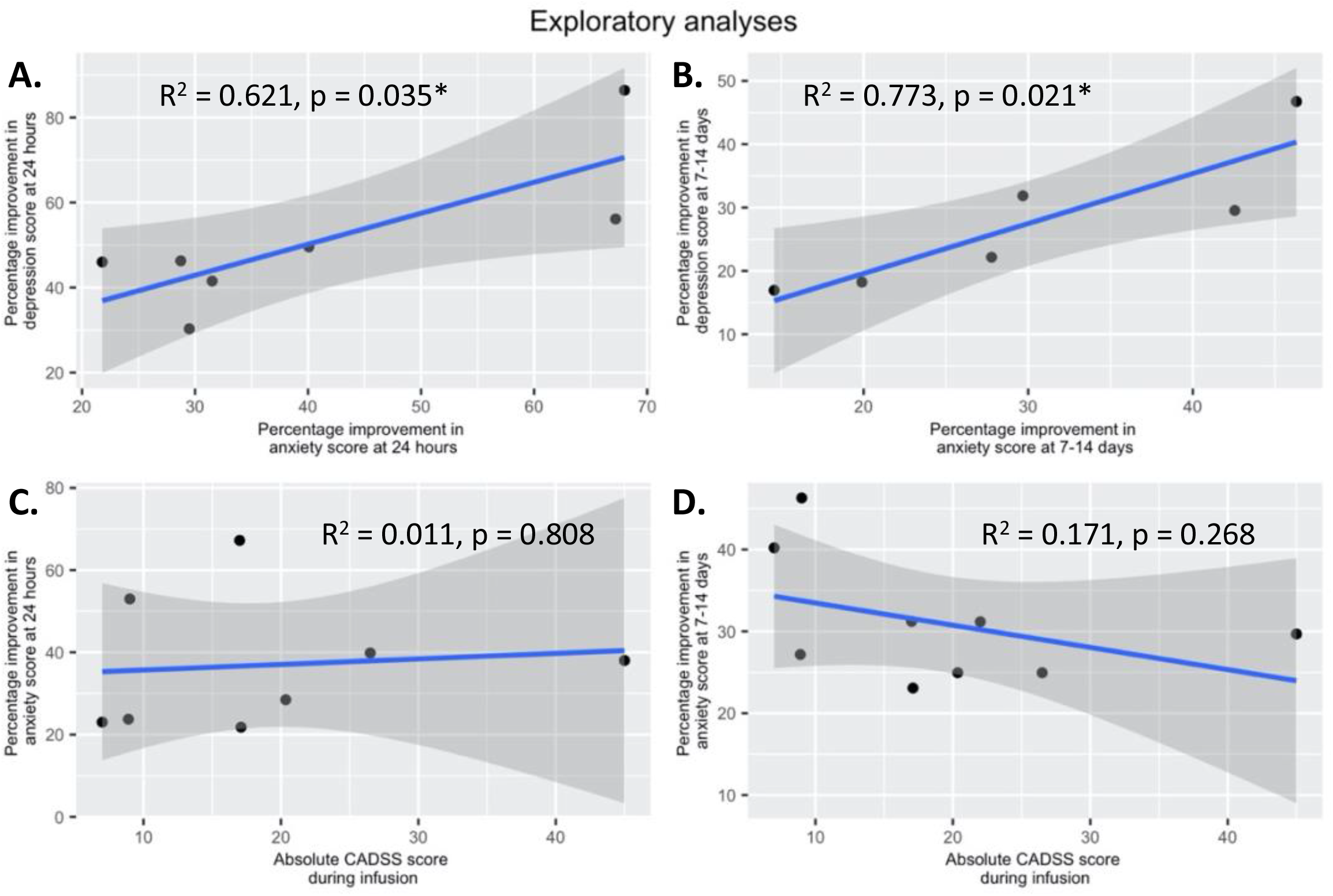
Exploratory analysis of the correlation between ketamine’s (**A**) anxiolytic and antidepressant effects at 24 hours (significant positive correlation; R^2^ = 0.621, *p* = 0.035); (**B**) anxiolytic and antidepressant effects at 7-14 days (significant positive correlation; R^2^ = 0.773, *p* = 0.021); (**C**) anxiolytic effects at 24 hours and peak Clinician-Administered Dissociative States Scale (CADSS) scores (no correlation; R^2^ = 0.011, *p* = 0.808); (**D**) anxiolytic effects at 7-14 days and peak CADSS scores (no correlation; R^2^ = 0.171, *p* = 0.268).

### Correlation between improvement in anxiety and peak dissociation

We sought to explore the relationship between peak dissociation and anxiety scores by correlating peak Clinician-Administered Dissociative States Scale (CADSS; Bremner et al., 1998) scores with percentage improvement in anxiety at the subacute and sustained time points. All peak CADSS scores were during infusion, except for Taylor et al. (2018), which was measured at one-hour post-ketamine administration.

At 24 hours post-administration, data from 222 participants was pooled to be included in the analysis. Results revealed no significant correlation between peak level of dissociation and improvement in anxiety symptoms (R^2^ = 0.011, *p* = 0.808; **Figure 4C**). Similarly, at 7-14 days post-administration, where data from 237 participants were pooled, there was no significant correlation between the two scores (R^2^ = 0.171, *p* = 0.268; **Figure 4D**).

## Discussion

This is the first transdiagnostic systematic review and meta-analysis of RCTs to assess the temporal profile of ketamine’s anxiolytic effects after a single dose. Of the 14 studies included in the systematic review and meta-analysis, seven studies assessed ketamine’s anxiolytic effects in PTSD; five in mood disorders; two in anxiety disorders; and two in either chronic or cancer-related pain. Results suggest that ketamine is an efficacious treatment for anxiety symptoms across this range of settings. Ketamine’s anxiolytic effect typically emerged after three to four hours and continued to be significantly superior relative to placebo at 24 hours and 7-14 days post-administration. Our findings suggest that single ketamine infusions could therefore offer both rapid and sustained improvement in anxiety. This work builds on previous systematic reviews and meta-analyses supporting ketamine’s use in anxiety disorders (Banov et al., 2020; Tully et al., 2022; Whittaker et al., 2021).

Seven studies reported data <12 hours after administration. Our results at this acute time point support the conclusions made by both preclinical and clinical studies that ketamine may result in rapid reductions in anxiety (Riehl et al., 2011; Glue et al., 2017). The earliest significant effect was reported by Zarate and colleagues (2012) at 40 minutes post-infusion with the majority of studies measuring anxiety symptoms at this time point reporting a significant effect at three to four hours. This corroborates the finding by Glue and colleagues (2017) in an open-label study, where ketamine administration led to marked improvements in anxiety scores at one-hour post-dose in patients with generalised anxiety disorder and/or social anxiety disorder.

10 studies reported data at 24 hours post-administration – the modal time point in the meta-analysis – where ketamine again had a beneficial effect compared to placebo. This aligns with results from responder data, which provides an additional insight into whether a drug provides clinically meaningful symptom relief. Glue and colleagues showed that patients with SAD and GAD show a higher likelihood of response at 24 hours after ketamine compared to midazolam (Glue et al., 2020).

Nine studies reported data at 7-14 days after administration. The sustained time point had the most RCTs report significant drug differences out of all time points. This suggests that ketamine has anxiolytic effects which last through one to two weeks post-treatment. As such, our findings closely match the literature exploring the drug’s efficacy in depression, which has found ketamine to have an anti-depressant effect lasting 7-14 days after a single dose (Coyle & Laws, 2015)

We endeavoured to examine any anxiolytic effects beyond 14 days. However, very few studies met our inclusion criteria (*n* = 4). From the included studies, there is evidence that ketamine continues to be superior to placebo beyond two weeks post-administration, with one study finding evidence of an anxiolytic effect up to one-month after a single dose (Pradhan et al., 2018). Coupling these results with the optimism in findings from earlier time points, future studies should further explore ketamine’s sustained anxiolytic efficacy beyond that of two weeks.

To date, most research into the psychiatric utility of ketamine has focused on depression. We conducted an exploratory analysis into the relationship between depression and anxiety improvements at both the subacute and sustained time point. If the anxiolytic and antidepressant effects of ketamine share similar mechanisms, one might expect to see a strong correlation between the improvements in both symptoms. Results revealed a significant correlation between mean percentage improvements in depression and anxiety at both the subacute (*k* = 7) and sustained time points (*k* = 6). Future imaging work could explore whether the same or distinct neural circuits which are related to ketamine’s effects on distinct symptom clusters.

We also analysed whether there was any link between ketamine’s anxiolytic effect and its peak dissociative effects as measured using the CADSS, at both the subacute and sustained time points. Research on the relationship between dissociation and therapeutic outcomes of ketamine treatment is equivocal. There are documented concerns that ketamine’s dissociative effects may negatively impact anxiety outcomes (Carlson et al., 2012; Coutinho et al., 2016; Schönberg et al., 2005 & 2008). Conversely, there is the suggestion that the degree of dissociation is important in ketamine’s therapeutic effects, and specifically that greater levels of dissociation may lead to more improvement in depression symptoms (Correia-Melo et al., 2017; Luckenbaugh et al., 2014; Sos et al., 2013). Our analysis showed no significant relationship between the two at either time point, suggesting that ketamine’s anxiolytic effects are independent from its dissociative effects. Indeed, there is research which has also concluded no relationship between peak dissociation and therapeutic effect (Berman et al., 2000; Ballard & Zarate, 2020; Fava et al., 2020; Lapidus et al., 2014; Włodarczyk et al., 2021). Nevertheless, our analysis was done using a small number of data points and more work should be done to further investigate these conclusions.

Several limitations of our study are of note. First is the high risk of bias in most of the included studies. Nine out of the 11 studies were reported as having high risk of bias, mainly because of unblinding of patients and outcome assessors, or selective reporting of data from secondary measures. The prevalence of unblinding in the included studies speaks to the difficulty in achieving effective blinding in studies exploring ketamine’s efficacy. Only five studies included an active placebo, and future research would benefit from the use of active placebos as controls. A second limitation is the presence of moderate heterogeneity in the meta-analysis of data at the acute time point (I^2^ = 50%) which implies clinical and/or methodological diversity in the included studies and implicates our conclusions. This may be due to low sample sizes (Inthount et al., 2015), and differences in study protocols (including the precise time point assessed within these windows) and disorders (though the latter is inherent in prioritising a transdiagnostic approach). While heterogeneity was low in the other two time points (I^2^ = 0% in subacute and sustained analyses), the potential for issues in comparability should not be ignored. Future work should aim for larger sample sizes and to carry out sensitivity analyses exploring the influence of diversity. Third, our meta-analyses consisted of findings from parallel arm and crossover studies. When carryover effects were found in crossover trials, data were limited to exclusively the first phase of the study; if not, collapsed data from both phases were used. It is possible that this variability introduced bias into our analysis. Finally, although this review intended to include studies assessing ketamine’s anxiolytic efficacy in settings such as chronic pain and palliative care, only one study in this context was eligible for inclusion (Dadabayev et al., 2020).

## Conclusion

Our meta-analysis demonstrates ketamine’s anxiolytic effects emerged rapidly at three to four hours post-administration and persisted for up to two weeks. This study is the first review to consider the efficacy of ketamine in anxiety symptoms across multiple time points using a transdiagnostic approach. By limiting our review to randomised control trials, which is considered as the highest quality of evidence (McErlean et al., 2022), we were able to eliminate any inconsistency due to methodological variation and identify areas of true uncertainty in the literature. Future RCTs should acknowledge that, in clinical settings, ketamine is most often administered in multiple doses and should explore anxiolytic effects after repeated dosing, as well as replicate our work to continue to elucidate the medication’s time course of action. This could help to identify the best administration pattern.

## Data Availability

All data produced in the present study are available upon reasonable request to the authors

## Author contributions

Conceptualisation, A.H.Y., L.A.; Methodology, H.H., K.M., L.A.J., R.S., A.H.Y., L.A.; Formal analysis, H.H., K.M., R.S., L.A.; Investigation, H.H., K.M., L.A.; Writing – original draft, H.H., K.M., L.A.; Writing – review and editing, H.H., K.M., L.A.J., R.S., A.H.Y., L.A.; Visualisation, H.H., K.M., L.A.; Funding acquisition, A.H.Y.; Resources, A.H.Y.; Supervision, A.H.Y., L.A.

## Funding statement

This report represents independent research funded by the National Institute for Health Research (NIHR) Biomedical Research Centre at South London and Maudsley NHS Foundation Trust and King’s College London. The views expressed are those of the authors and not necessarily those of the NHS, the NIHR, or the Department of Health. Dr Luke A Jelen is a Medical Research Council (MRC) Clinical Research Training Fellow (MR/T028084/1). Dr Laith Alexander is an NIHR Academic Clinical Fellow in Translational Psychiatry (ACF-2022-17-016).

## Declaration of interests

HH, KM, LJ and LA have no conflicts of interest. In the last 3 years: RS declares an honorarium from Lundbeck. AHY declares honoraria for speaking from Astra Zeneca, Lundbeck, Eli Lilly, Sunovion; honoraria for consulting from Allergan, Livanova and Lundbeck, Sunovion, Janssen; and research grant support from Janssen.

## Notes

### Clinical Protocols

https://www.crd.york.ac.uk/prospero/display_record.php?ID=CRD42022303070

