## Supplementary for "A transdiagnostic systematic review and meta-analysis of ketamine’s anxiolytic effects"

| Lead author | Background | Findings reported by the authors |
|---|---|---|
| Abdallah et al. (2022) | Abdallah et al. (2022) randomly assigned 158 participants with PTSD to one of three groups: saline placebo, low dose ketamine (0.2 mg/kg), and standard dose ketamine (0.5 mg/kg). Infusions were given twice weekly for four weeks. | **Acute:** NA |
| | | **Subacute:** Significant reduction in scores using the PCL-5 between ketamine and placebo groups, which became non-significant when adjusting for multiple comparisons (mean difference [± SEM] from placebo = 6.6 [±3.1], $t_{149}$ (149) = 2.1, $p = 0.04$, adjusted $p = 0.11$). |
| | | **Sustained:** Authors collected PCL-5 scores at seven, 11- and 14-days post-ketamine. Results revealed no significant drug group differences between seven and 14 days after treatment began, and authors reported no significant treatment-by-time interaction ($F_{18,137} = 1.1$, $p = 0.38$) or main effect of drug. There was a significant time effect on PCL-5 scores ($F_{9,133} = 37.1$, $p < 0.0001$), but it was present in all study participants |
| | | **Beyond 14 days:** NA |
| Dadabayev et al. (2020) | Assessed the efficacy of ketamine compared to ketorolac in reducing PTSD symptoms in 41 chronic pain patients with and without PTSD. | **Acute:** NA |
| | | **Subacute:** PTSD symptoms were significantly reduced at 24 hours compared to baseline ($t_{32.59} = 2.33$, $p = 0.03$) in all participants, so there were no significant differences between the two medication groups ($p > 0.05$). |

| | | |
|---|---|---|
| | | **Sustained:** Authors reported a significant decrease in IES-R scores from baseline to day 7 in both the ketamine and ketorolac groups ($F_{1,52}$ = 9.35, $p < 0.01$), but no significant drug group differences. |
| | | **Beyond 14 days:** NA |
| Fallon et al. (2018) | Administered either oral ketamine or oral inert placebo to patients with cancer-related neuropathic pain (n=214). The ketamine and placebo doses were titrated over the course of 2 weeks, and the stable dose was then continued to be administered for 16 additional days. | **Acute:** NA |
| | | **Subacute:** NA |
| | | **Sustained:** NA |
| | | **Beyond 14 days:** Analysis using the Hospital Anxiety and Depression Scale (HADS; Zigmond & Snaith, 1983) revealed a non-significant mean difference between the two treatment groups of 0.020 (95% CI: [-0.500, 9.250]) with an adjusted AUC of 0.92. |
| Feder et al. (2014) | Compared the efficacy of 0.5mg/kg ketamine with an active placebo, midazolam (0.045mg//kg), in 41 patients with chronic PTSD. The order of infusions was randomly assigned, and a two-week washout period was used between infusions, which successfully prevented carryover effects. | **Acute:** NA |
| | | **Subacute:** PTSD scores had significantly improved in those receiving ketamine compared to those receiving midazolam with a group mean difference of 12.7 ([95% CI: 2.5 - 22.8]; $p = 0.02$). |
| | | **Sustained:** Authors analysed IES-R scores at seven days but did not report significance for this time point. However, they reported their |

| | | |
|---|---|---|
| | | secondary measure (CAPS scores) and there was no significant difference in mean CAPS scores between treatment groups (mean difference between ketamine and midazolam: 8.7 [95% CI, -4.8 to 22.2]; *p* = 0.20). |
| | | **Beyond 14 days:** NA |
| Feder et al. (2021) | Assessed the sustained effects of ketamine in a sample of chronic PTSD patients (*n* = 30). Using a parallel arm design, participants were randomised to receive six infusions of either 0.5mg/kg intravenous ketamine or 0.045 mg/kg intravenous midazolam in rapid succession over two weeks (three per week). Scores from the Clinician-Administered PTSD Scale for DSM-5 (CAPS-5; Weathers et al., 2018) were used to assess symptom severity one week into treatment (before fourth infusion) and at the end of treatment. | **Acute:** NA |
| | | **Subacute:** Reported non-significant findings regarding total IES-R scores |
| | | **Sustained:** Analysis revealed significantly lower total scores in the ketamine group compared to the midazolam group at one week post-first infusion (estimated difference= 8.80, SE = 3.93, *p* = .030), which was sustained at two-weeks post-first infusion (estimated difference= 11.88, SE = 3.96, *p* = .004). |
| | | **Beyond 14 days:** NA |
| | | **Acute:** Did not report findings |

| | | |
|---|---|---|
| Lapidus et al. (2014) | Administered 50mg intranasal ketamine or saline solution to 20 patients with major depressive disorder (MDD). Assessed anxiety using HAM-A scale. | **Subacute:** Ketamine significantly reduced scores on the HAM-A scale at 24 hours compared to placebo ($t_{17} = 3.06$, $p = 0.007$; mean benefit of 4.5 points [95% CI: 1.4 – 7.6]). |
| | | **Sustained:** NA |
| | | **Beyond 14 days:** NA |
| Murrough et al. (2015) | Administered participants with either an anxiety ($n = 12$) or mood ($n = 12$) disorder, with clinically significant suicidal ideation, either 0.5 mg/kg IV ketamine or 0.045 mg/kg IV midazolam over 40 minutes. | **Acute:** NA |
| | | **Subacute:** Patients randomly allocated to the ketamine arm did not significantly differ in their CAST anxiety subscale ratings from those in the control arm ($F_{1,21} = 1.74$, $p = 0.20$). |
| | | **Sustained:** Authors did not analyse the CAST subscale scores at seven days post-infusion, though group level data from the CAST anxiety subscale was included in the meta-analysis. On the subscales pertaining to anxiety (irritability, anxiety and panic), the ketamine group had numerically lower mean scores than the midazolam group on each subscale. |
| | | **Beyond 14 days:** NA |
| Norbury et al. (2021) | A clinical trial of repeated-dose intravenous ketamine (0.5mg/kg) compared to midazolam | **Acute:** NA |
| | | **Subacute:** NA |

| | | |
|---|---|---|
| | (0.045mg/kg) in 21 participants with PTSD. Participants had three infusions per week for two weeks | **Sustained:** NA<br><br>**Beyond 14 days:** CAPS-5 scores were significantly lower at around 16 days post-first dose in patients who had received ketamine than those who had received midazolam. A repeated-measures ANOVA with drug, session, and measure factors revealed a significant session-by-drug interaction on CAPS-5 and MADRS scores ($F_{1,57}$ = 6.58, $p$ = 0.013). |
| Nugent et al. (2019) | A cross-over study administering 0.5 mg/kg intravenous ketamine and intravenous saline solution to 35 subjects with treatment-resistant MDD and 25 healthy controls. The aim of the study was broadly to help delineate ketamine's mechanism of action in TRD patients, and to observe ketamine's effects on mood and EEG-measured resting gamma power.<br>The HAM-A was administered to measure changes in anxiety symptoms at 40 and 230 minutes as well as at one-, two- and three-days post-infusion. | **Acute:** Results revealed a main effect of both drug ($F_{1,101}$ = 38, $p$ <0.001) and time ($F_{9,110}$ = 5.2, $p$ < 0.001), with no significant drug-by- time interaction ($F_{9,126}$ = 0.9, $p$ = 0.51). In MDD patients alone, there was no significant change in anxiety symptoms at 40 minutes post-ketamine compared to placebo (estimated Cohen's d: 0.00, 95% CI: [-0.34, 0.34]), but at 230 minutes, authors reported that those receiving ketamine showed a significantly greater reduction in anxiety scores than those who received placebo (estimated Cohen's d: 0.60, 95% CI: [0.26, 0.94]).<br><br>**Subacute:** MDD patients demonstrated significantly greater reductions in HAM-A scores at 24 hours after receiving intravenous ketamine compared to placebo (estimated Cohen's d: 0.58 [95% CI: 0.24, 0.92]). |

| | | |
|---|---|---|
| | | **Sustained:** No significant differences between those who received ketamine and those who received placebo (seven days: group mean difference 2.45 [95% CI: -1.276, 6.176]; 10 days: 1.98 [95% CI: -1.021, 4.981]). |
| | | **Beyond 14 days:** NA |
| Pradhan et al. (2017) | Crossover clinical study where 10 chronic and refractory PTSD patients were randomly allocated to one of two groups. The first group received 12 sessions of Mindfulness Based Extinction and Reconsolidation (TIMBER) psychotherapy alongside a single intravenous infusion of ketamine (0.5mg/kg), whilst the second group underwent the same TIMBER psychotherapy sessions but instead received a saline infusion. Participants in the placebo condition were switched to the ketamine infusion group once they had experienced sustained relapse for two weeks. | **Acute:** There was a significant decrease from baseline in PCL scores at 4 hours post-infusion across all participants ($p$ = 0.003), with no significant difference between the groups of the study ($M_{ketamine}$ = 37.80, $SD_{ketamine}$ = 20.34; $M_{placebo}$ = 55.20, $SD_{placebo}$ = 13.31, $p$ = 0.148). The CAPS was not administered at this time point. |
| | | **Subacute:** Reported no significant differences on the PCL or CAPS measures between the two arms of their study at 24 hours ($M_{PCL\ ketamine}$ = 25.60, $SD_{PCL\ ketamine}$ = 3.78, $M_{PCL\ saline}$= 26.60, $SD_{PCL\ saline}$ = 7.63, $p$ = 0.80; $M_{CAPS\ ketamine}$ = 17.80, $SD_{CAPS\ ketamine}$ = 5.21, $M_{CAPS\ saline}$= 23.40, $SD_{CAPS\ saline}$ = 8.99, $p$ = 0.26), although both groups demonstrated significant reductions from baseline on both the PCL ($p$ <0.001) and the CAPS ($p$ < 0.001). |
| | | **Sustained:** NA |

| | | |
|---|---|---|
| | | **Beyond 14 days:** Patients receiving ketamine had a more sustained response (33 ± 22.98 days) than those who received placebo (25 ± 16.8 days), though this difference was not significant (*p* = 0.545). |
| Pradhan et al. (2018) | Randomised a sample of 20 patients (different to those recruited in the 2017 study) with refractory PTSD to receive either 0.5 mg/kg IV ketamine over 40 minutes or IV saline. Both groups received 12 sessions of TIMBER psychotherapy alongside either a single intravenous infusion of ketamine (0.5mg/kg) or saline. | **Acute:** NA |
| | | **Subacute:** No significant between group differences in PCL and CAPS scores at 24 hours post-infusion, but instead both participant groups experienced a significant reduction in PTSD symptoms on both measures ($p < 0.0001$). |
| | | **Sustained:** NA |
| | | **Beyond 14 days:** Patients receiving ketamine had a significantly more sustained response (34.44 ± 19.12 days) compared to those who received placebo (16.50 ± 11.39) ($p = 0.022$). |
| Rodriguez et al. (2013) | Assessed the effects of a 40 minute 0.5mg/kg intravenous ketamine infusion compared to a 0.9% saline infusion in fifteen medication-free OCD patients who experienced near-constant obsessions. | **Acute:** Subjects who had been given ketamine had significantly lower scores than those who received placebo on the OCD-VAS at 230 minutes post-infusion. Specifically, the mean score of the ketamine group was 3.84 points lower (SE = 1.59, $p < 0.05$) than the mean of the placebo group. The OCD-VAS was also administered at 90 minutes and 110 minutes post-infusion, but no significant findings were reported at either time point. |

| | | |
|---|---|---|
| | Note: A one-week washout period was used before participants switched solution however, significant carryover effects were identified so data could not be collapsed across phases. Thus, only data from the first phase of the study were used to assess outcome measures. | **Subacute:** No significant differences in anxiety scores between the ketamine and saline conditions.<br>**Sustained:** There were significantly lower means on the OCD-VAS at seven days post-infusion in subjects who had received IV ketamine compared to those who had received placebo (by -3.67, SE = 1.36, $p < 0.05$)<br>**Beyond 14 days:** NA |
| Taylor et al. (2018) | Studied adults (*n* = 18) with a DSM-5 diagnosis of social anxiety disorder in a double-blind, randomised controlled crossover trial. Participants were assigned to initially receive either 0.5 mg/kg intravenous ketamine over 40 minutes or intravenous saline before receiving the alternate dose 28-days later. Significant carryover effects on the LSAS but not on the VAS scale. | **Acute:** Found no significant differences between ketamine and placebo groups on primary anxiety outcome measures at the 3-hour time point and did not report any significant change from baseline (statistics not reported).<br>**Subacute:** Found no significant improvements in LSAS scores at 24 hours although statistical tests results were not reported.<br>**Sustained:** Found a significant time × treatment interaction on the LSAS (Time × Treatment: $F_{9,115}= 2.6$, $p = 0.001$). Specifically, the authors report significant between-group differences emerging at day 10 post-infusion, with the ketamine group demonstrating significant reductions in overall LSAS scores compared to placebo counterparts (difference = 28.72 ± 7.36, ES = 1.37, $F = 7.6$, $p = 0.01$). |

| | | |
|---|---|---|
| | | **Beyond 14 days:** NA |
| Zarate et al., (2012) | Assessed ketamine's effect on Hamilton Anxiety Scale (HAM-A; Hamilton, 1959) scores and Anxiety Visual Analogue Scale (VAS-Anxiety; Aitken, 1969) scores in a sample of 15 bipolar disorder subjects. All participants were maintained on therapeutic levels of lithium or valproate before being randomised to receive either a single infusion of ketamine (0.5mg/kg) or saline. Subjects were blindly administered two infusions with a two-week interval between each session. | **Acute:** A significant drug-by-time interactions for both measures (HAM-A: $F_{7,141}$ = 2.75, $p$ = 0.01; VAS-Anxiety scale: $F_{10,166}$ = 2.12, $p$ = 0.03) with no carryover effects between placebo/ketamine phases. Specifically, ketamine decreased symptoms of anxiety significantly more than placebo starting at 40 minutes post-infusion and remained significant at 80 minutes, 110 minutes, and 230 minutes (HAM-A was only significantly different at 230 minutes). |
| | | **Subacute:** Found ketamine to be superior to placebo at symptom reduction using HAM-A at 24 hours ($F_{7,141}$ = 2.75, $p$ = 0.01). |
| | | **Sustained:** Reported significantly lower scores in subjects who had received ketamine as opposed to placebo on the VAS-Anxiety scale at days seven and 14 post-ketamine administration ($F_{10,166}$ = 2.12, $p$ = .03). |
| | | **Beyond 14 days:** NA |

**Supplementary Table 1.** A detailed summary of significant and non-significant findings of each study.
